# Current Attitudes and Use Patterns of Large Language Models in Clinical Practice and Medical Education Among Trauma Providers

**DOI:** 10.64898/2026.10.20.26366047

**Authors:** Brendon Frankel, Chavi Rehani, Elizabeth Anne Clark, Susan L. Evans, Vicki Moran, Hannah K. Galvin, Lizette Villarreal, Andrea L. Nevedal, Ashley N. Moreno, Mohamed Albirair, Lacey N. LaGrone, The 2026 Design for Implementation (DFI) Authorship Group, The Design for Implementation (DFI) 2026 Authorship Group, Al B. Benson, Pamela Bixby, Stephanie Bonne, Eileen M. Bulger, Jennifer Marie Chastek, Chelsea Church, Elizabeth Anne Clark, Todd W. Costantini, Nicholas Paul Cozzi, Kimberly A. Davis, Tomas Drabek, Brian John Eastridge, Evert Austin Eriksson, Susan Lynn Evans, Shannon Marie Foster, Brendon Mark Frankel, Hannah K. Galvin, Jasmine Garces-King, Jeffrey Michael Goodloe, Jenny M. Guido, Katheryn Grider, Jennifer L. Hartwell, Elliott R. Haut, Laura Sabine Johnson, Alyssa B. Johnson, Suren Kanayan, Christopher Scott Kang, Anai N. Kothari, Morgan Ashley Krause, Kelly L. Lang, Jessica Marie Lee, Philipp Lirk, Halinder S. Mangat, Debra M. Marvel, Christopher P. Michetti, M. Victoria P. Miles, Charles Mock, Vicki Moran, Ashley N. Moreno, Abirami Muthumani, Priyansh Prakash Nathani, Nobhojit Roy, Katherine Theresa Flynn-O’Brien, Jennifer Anne Pekarek, Michael Allen Person, John Kenneth Petty, Michelle A. Price, Bradford L. Ray, Diane Redmond, Chavi Rehani, Linda Kate Reinhart, Shannon Rosenauer, Rowan Rice, Babak Sarani, Sebastian D. Schubl, Erica Shelton, Jonathan Daniel Stallings, Deborah M. Stein, Erik G. Van Eaton, Cynthia Lizette Villarreal, Jennifer J. Ward, Jeffrey Lee Wells, James “Tripp” E. Winslow, Lydia M. Wood, Arnold D. Wynn

**Author notes:** **CORRESPONDING AUTHOR**: Lacey N. LaGrone, MD, MPH, MA, 2500 Rocky Mtn Ave; Loveland, CO 80525, Trauma Acute Care Surgery, Medical Center of the Rockies, Loveland, CO, USA UCHealth; University of Colorado.

## Abstract

**Introduction:** Large language models (LLMs) are increasingly used in clinical practice and medical education, yet uncertainty remains regarding appropriate use, oversight, and potential effects on clinical reasoning and patient safety. This study examined current LLM use patterns, attitudes, perceived risks, and priorities for responsible integration among individuals engaged in trauma care, clinical guidance development, and medical education.

**Methods:** Participants attending the third Design for Implementation: The Future of Trauma Clinical Guidance and Research Conference (DFI) completed surveys examining LLM use, attitudes, perceived inhibitors to personal use, and clinical applications. During facilitated sessions, participants used open-ended note-taking guides to document their perspectives regarding LLM use in clinical practice, education, and guidance development. Survey data were analyzed descriptively, and qualitative responses were analyzed using rapid qualitative content analysis.

**Results:** 77 participants completed the pre-conference survey; 62 clinicians provided data regarding clinical LLM use and attitudes. Among clinicians, 15% reported daily LLM use, whereas 26% reported never using LLMs. Respondents expressed strongest agreement with the need for regular quality auditing of LLM-supported clinical decisions. Among 52 post-conference survey respondents, 96% somewhat or strongly agreed that clinical training curricula should include education on LLM use. Barriers to LLM use included time required to verify accuracy, concern about patient harm, and insufficient training in appropriate prompting. Qualitative data came from 23 note-taking documents administered during the conference. Both positive and negative perspectives arose regarding AI use, centering on topics including efficiency gains, deskilling and never-skilling, governance and oversight, accuracy and hallucination, patient safety, and education.

**Discussion:** DFI participants recognized efficiency and informational benefits of LLMs while identifying important concerns related to accuracy, patient safety, skill development, and governance. Findings support a need for institutional guidance, education in responsible LLM use, and evaluation of the incorporation of these technologies into clinical practice and health professions education.

**What is already known on this topic:** LLMs have become prevalent across professional domains in recent years, including medical education and clinical practice.

**What this study adds:** Through interactive sessions, key stakeholders working predominantly at urban academic medical centers characterized the current ways in which they see LLMs being used most frequently and the concerns they have with this use.

**How this study might affect research, practice, or policy:** This study demonstrates a need to further understand current use patterns of LLMs in medical education and clinical practice, which would be necessary to develop guidelines for use and/or the oversight that many stakeholders identified as necessary.

## Introduction

Since the public launch of ChatGPT in November 2022, artificial intelligence (AI) and large language models (LLMs), which use AI to generate human-like responses to written prompts, have been subjects of frequent discussion and debate. Systems such as ChatGPT, Claude, and Gemini have been adopted to complete personal and professional tasks more efficiently; however, there is a lack of consensus around the appropriate application of these tools for the purposes of medical education and decision-making in clinical practice. Despite this lack of consensus, these tools have become common in clinical use. For example, a recent survey of emergency physicians from the American College of Emergency Physicians found that over half (61%) of emergency physicians and residents reported at least some use of AI.^1^ Their use in trauma care is equally important.^2^

Beyond tools such as ChatGPT that can be used regardless of subject matter, a subset of LLMs known as retrieval-augmented generation (RAGs) that pull information from specific types of medical literature have gained popularity among medical providers.^3,4^ These programs may be useful in synthesizing well-established, curated, peer-reviewed medical knowledge as they improve, and they can vastly increase efficiency in researching a particular medical topic. One RAGs’ recent 100% accuracy score on the United States Medical Licensing Examination (USMLE) and gradually higher test scores with each ChatGPT iteration demonstrate these improvements.^5,6^

Medical specialty clinical knowledge tools, many of which now have HIPAA-compliant features including Electronic Health Record integration capabilities for context-aware searching, all show promise.^7^ However, many clinicians and educators have cited concerns about use of these programs at early stages of medical education and impacts on training due to possible effects on younger trainees. “Never-skilling” describes the concern that over-reliance on LLMs may adversely impact the development of clinical reasoning skills that have long been viewed as essential to safe and independent medical practice.^8^ Another term, “de-skilling,” has been used to describe an analogous concern related to the loss of independent decision-making in currently practicing physicians and surgeons, whereas “automation bias” refers to a clinician’s predisposition to trust the output of such tools.^3,9,10,11^ A landmark Lancet observational study on de-skilling found that after even brief use of AI-assisted colonoscopies among endoscopists in Poland, the same physicians’ adenoma detection rate without AI significantly decreased in subsequent colonoscopies, demonstrating possible over-reliance on these technologies once introduced.^12^

Furthermore, these medical specialty models are specifically designed to retrieve information from open access medical literature and journals or organizations with which they specifically license content. ^5^ This may reduce the likelihood of hallucinations and increase transparency.^4^ However, these tools may only be able to access article abstracts beyond these sources instead of full articles, and recent studies have shown mixed results on their accuracy; while some studies show that LLMs and RAGs specific to medicine are more accurate and develop safer management plans, other studies show that broad-based, foundational LLMs (i.e., not medicine-specific) outperform medical specialty _models._ 7,13,14

The result is an output of answers from these models that may be incomplete or inaccurate based on the well-documented phenomenon of AI hallucination when asked relatively ambiguous questions. At an early clinical stage of training, and in a rapidly evolving patient scenario requiring prompt decision-making with limited information, these constraints could pose a risk to patient safety and inhibit trainees’ skill development to rapidly process and appropriately act on information.

To date, several studies have quantified the prevalence of clinicians’ LLM use, but few have also described their attitudes toward using these tools for clinical guidance development and decision-making.^1,15,16^ The aims of this study are: 1) using quantitative methods, identify frequency of, and attitudes toward, using LLMs and AI in clinical decision-making among trauma clinicians engaged in national collaborative clinical guidance efforts; and 2) using qualitative methods, assess these clinicians’ attitudes about AI in clinical guidance including: oversight, impacts on trainee skill development, accuracy, education, values and patient safety.

## Methods

### Study design and research personnel

This IRB-approved (Colorado Multiple Institutional Review Board, CB F490; COMIRB No: 24-0047) mixed methods study took place at the third Design for Implementation: The Future of Trauma Research & Clinical Guidance (DFI) Conference, held at the American College of Surgeons (ACS) Headquarters in Chicago, Illinois in February 2026. Open-ended responses and ordinal survey data were used to identify current patterns of LLM/AI use among conference attendees in clinical decision-making. Qualitative, free-text data from a self-administered, structured note-taking tool were used to assess attendees’ priorities for responsible LLM use across several domains including oversight, impacts on trainee skill development, accuracy, education, values and safety.

The research team included a medical student (BF), two trauma surgeons (LNL, SE), a program manager (ANM), two research design consultants (AN, MA), two medical residents (CR, EC), a registered nurse (VM), a pediatrician and subject matter expert on AI use in medicine (HG), and four pro bono management consultants (SR, CC, RR, KG). Data were collected from February 25-26, 2026 by BF and ANM and analyzed primarily by BF. Facilitators for each discussion group (CR, SE, VM, EC) were selected based on expertise pertinent to the article topic and attended a two-hour overview and training session before the conference.

### Sample and procedures

Conference attendees were invited primarily via the network established by the first two years of the DFI conference and through the Coalition for National Trauma Research.^17,18^ All conference attendees were invited to take the pre- and post-survey and participate in discussion groups. Respondents were not required to have completed the pre-survey in order to complete the post-survey, nor to attend discussion groups. All three data collection activities were administered independently and were neither required nor mutually exclusive. Attendees self-selected to complete surveys and attend discussion groups. Informed consent was provided through the pre-conference survey. Participant confidentiality was prioritized throughout the study.

#### Survey procedures

Microsoft Forms (Microsoft 365, V2505) survey links were sent via email to all registered in-person and virtual attendees. The pre-conference survey was sent five days before the conference began. The survey was sent once, and no follow-up reminders were sent via email. A printed QR code was displayed at the registration table on the first day of the conference to encourage people to complete the survey if they had not yet done so. The post-conference survey was sent to all registrants the day after the conference concluded. The survey was sent once, and no follow-up reminders were sent via email. The Consensus-Based Checklist for Reporting of Survey Studies was used for the conference surveys (Supplemental Item 1).^19^

#### Discussion group procedures

Conference participants attended a 20-minute presentation entitled “AI Use in Clinical Decision Making – Current State, Limitations, Priorities for Clarification,” an overview of the current accuracy and usefulness of LLMs for clinical purposes as well as insights into some of their limitations.

Prior to the conference, conference coordinators conducted a literature review on a wide range of LLM-related topics in clinical decision-making and medical education and then selected four articles based on relevance, topic diversity, and publication recency. The four selected articles were: “Preserving clinical skills in the age of AI assistance,” “Large Language Model Influence on Diagnostic Reasoning: A Randomized Clinical Trial,” “Can large language models replace humans in systematic reviews? Evaluating GPT-4’s efficacy in screening and extracting data from peer-reviewed and grey literature in multiple languages,” and “Reliability of LLMs as medical assistants for the general public: a randomized preregistered study.”^20,21,22,23^ These articles were shared via email with participants who were encouraged to read the articles prior to the conference. On Day One of the conference and following the AI Use presentation, in-person participants were split into three groups while virtual participants remained in one virtual room. Participant groups were cohorted to maximize the different geographies, demographic characteristics, and career types within each group. Participant groups rotated sequentially between four 20-minute sessions to discuss each of the four journal articles. Each group facilitator presented a brief synopsis of an individual article and led a guided discussion; each facilitator met once with each of the four groups. By the end of this session, all participants had discussed all four articles within their respective cohorts.

### Data tools

All data were collected in English.

#### Pre-conference survey

The pre-survey asked about current AI and LLM use in clinical practice, and participants’ level of concern (or lack thereof) on AI use across various clinical scenarios. All respondents also answered demographic questions. Instruments were not pilot tested. See Supplemental Item 2 for a consolidated list of pre-conference survey questions.

#### Post-conference survey

A post-conference survey was sent to all virtual and in-person attendees assessing level of concern (or lack thereof) on AI use across various clinical scenarios. Demographic data were not collected in the post-conference survey. See Supplemental Item 3 for a consolidated list of post-conference survey questions.

#### Discussion group notes

One structured note-taking template was provided to each participant to capture their impressions and feedback throughout the rotations (for in-person attendees, the notes were completed using pen/paper while virtual attendees filled out electronic versions). The template included four main questions that could be answered regardless of the specific article being discussed at the time:

- 1. What did you learn, if anything, from this discussion/topic that will change how you use AI?
- 2. What do you see working well, or what concerns do you have about LLM use?
- 3. What have you observed as the biggest change in clinical practice due to AI?
- 4. How does the article/discussion inform how we responsibly integrate AI into clinical guidance development and dissemination?

Demographic data were not collected in the note-taking documents. At the conclusion of the conference, facilitators collected the note-taking documents and BF transcribed the responses into Microsoft Excel (Microsoft Corporation, 2018).

### Data analysis

#### Quantitative analysis

Microsoft Excel was used to compile, tabulate and analyze all survey data (Microsoft Corporation, 2018). The post-conference survey was not linked to the pre-conference survey and neither was linked to the discussion group notes.

#### Qualitative analysis

Using techniques informed by rapid directed qualitative content analysis, the lead researcher (BF) developed a Microsoft Excel matrix (Microsoft Corporation, 2018) by sorting all paraphrased responses into several content-related categories pertaining to LLM use in clinical education and practice.^24,25,26^ Step one: data from five respondents were reviewed by BF to inductively identify participants’ reflections on use of AI which were grouped into the following six domains: Value and Efficiency Gains from AI, Deskilling and Never-Skilling, Governance and Oversight, Accuracy and AI Hallucination, Patient Safety, and Education on AI Use. These domains transcended the four individual questions and were not isolated to certain questions. Step two: these six domains were each assigned a column in the Excel spreadsheet, where each row represented a respondent. Step 3: using these six domains, BF assigned the remaining responses to relevant columns to understand salient perspectives about LLM use; we assumed flexibility to add or delete content areas throughout the process, but no additional salient domains were identified. BF reviewed each response for salience, repetition, and negative cases and proceeded with grouping together by topic. To ensure rigor, another researcher (AN) reviewed the results and methods used to identify content areas and assign responses. Once all responses were aligned to domains, BF reviewed responses and wrote results summaries for each domain to reflect overarching patterns and range of responses.

## Results

### Pre-Conference Survey Findings

#### Demographics of Conference Attendees

Seventy-seven attendees completed the pre-conference survey (Table 1). Most respondents were clinicians, including physicians (62%), registered nurses (12%), and advanced practice providers (5%). Among respondents reporting a clinical discipline, trauma surgery (49%), general surgery (37%), and emergency medicine (18%) were most frequently represented. 75% of respondents reported working in a verified or designated trauma center; among these respondents, 84% worked in a Level I trauma center. 17% reported practicing or working in a rural setting. In addition, 75% reported participating in clinical guidance development, 68% conducted research, and 65% participated in dissemination science or quality improvement activities.

**Table 1.**
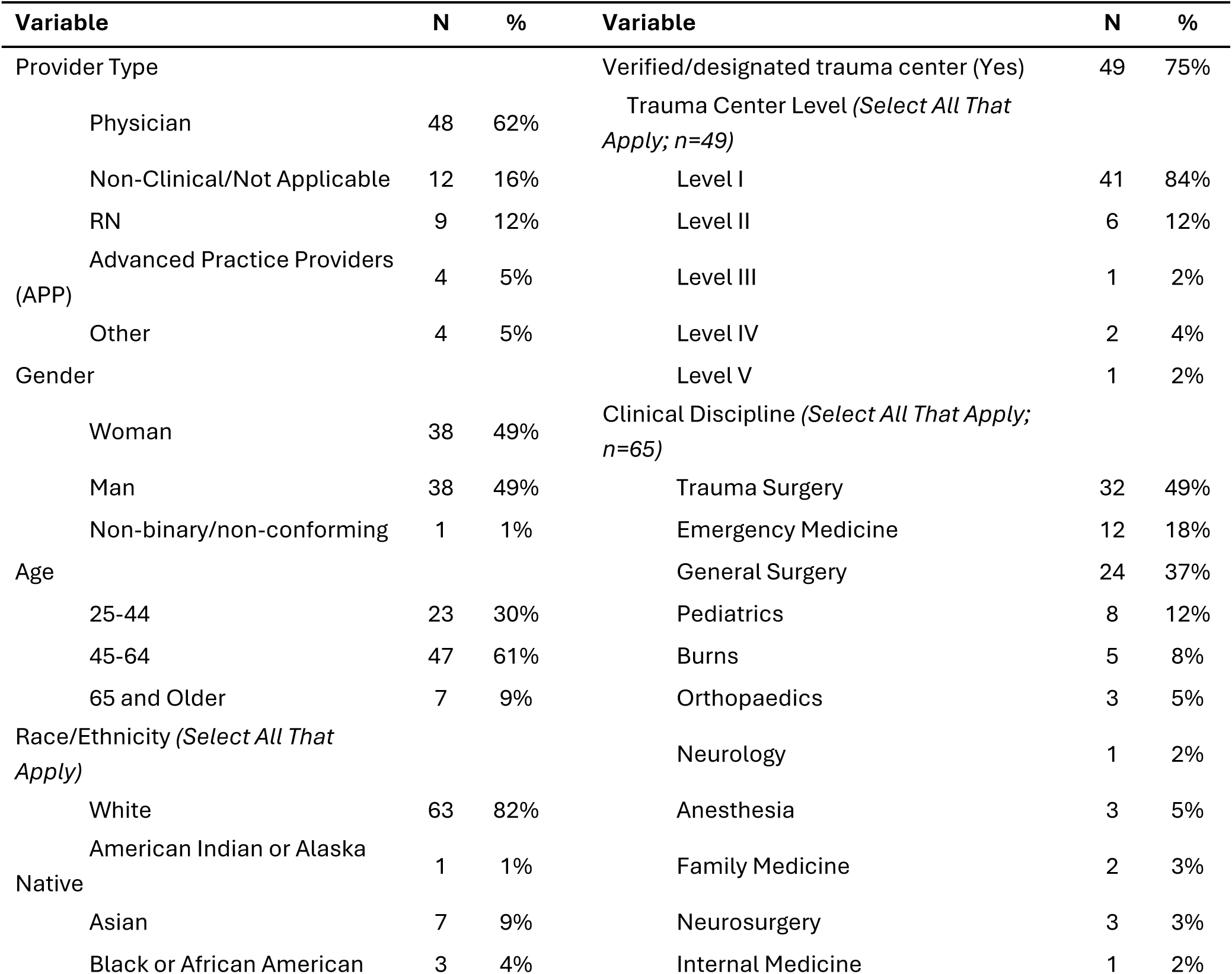
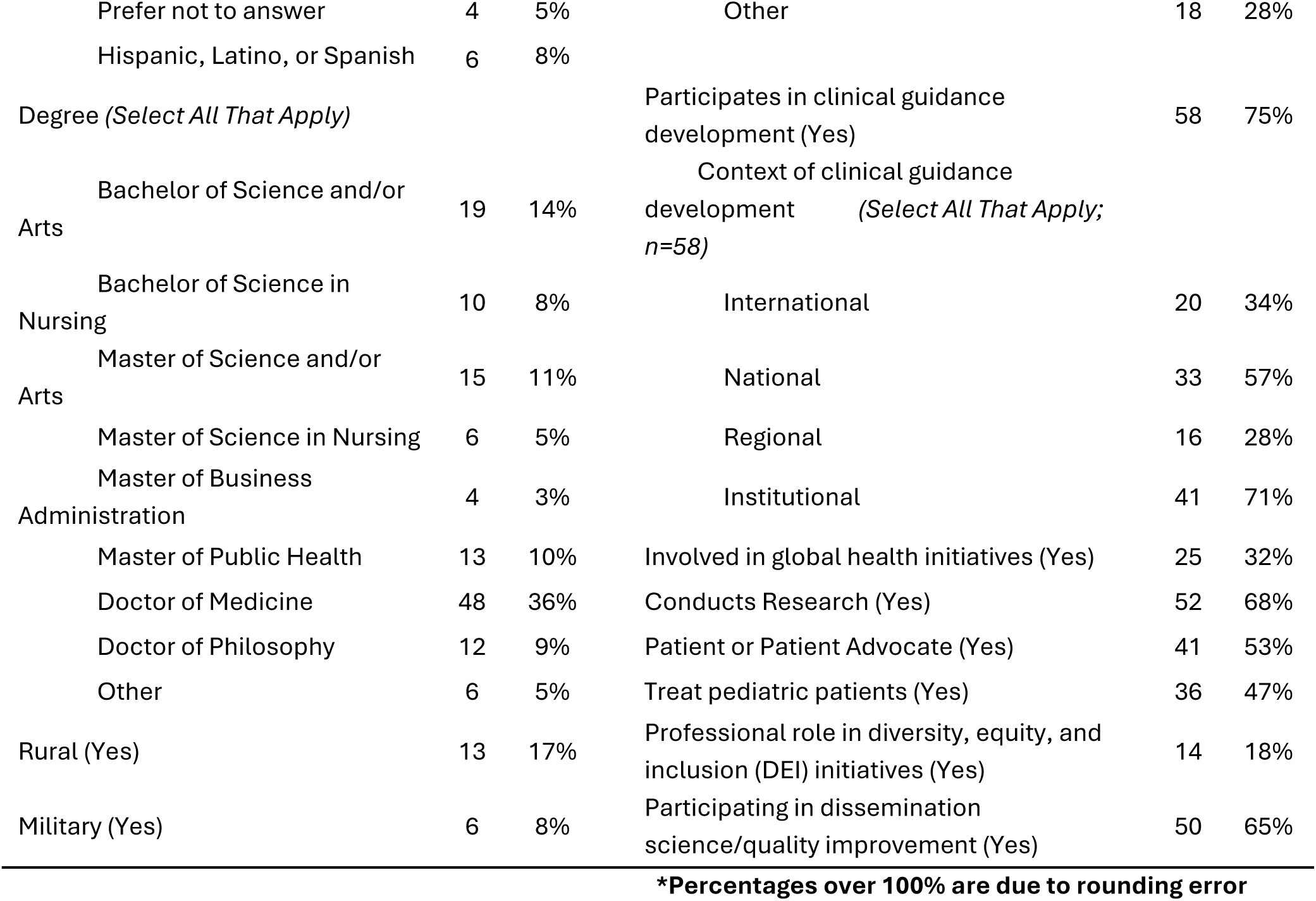
Demographics of Pre-Conference Survey Respondents (N=77)*

#### Use Patterns & Attitudes Towards LLMs

Only 5% of pre-conference survey responders identifying as clinicians cited LLMs or AI as the most frequently used information source when facing clinical uncertainty; peer-reviewed published literature (48%), peers and peer networks (24%), and summative point-of-care resources such as UpToDate (19%) were cited much more frequently.

General frequency of LLM use was also assessed among clinicians, with 26% of clinical responders indicating they never use LLMs and 15% indicating daily use. 41% of clinicians reported using AI or LLMs in very few clinical encounters, and only 4% indicated using them in most encounters. Table 2 shows rank-ordered responses to questions that assess attitudes towards AI and LLM use in clinical practice (on a scale of 1 = ‘strongly disagree’ up to 5 = ‘strongly agree’). The strongest agreement was expressed with statements regarding the need for quality control of decisions made with the help of LLMs, standardizing which LLMs can be used by clinicians, and the need to independently assess the validity of sources cited by LLMs.

**Table 2.** LLM Use and Attitudes of Pre-Conference Survey Respondents (N=62; clinicians only)

| **Question** | **Mean** | **SD** |
| --- | --- | --- |
| LLMs used to support clinical decisions should be regularly audited for quality control | 4.53 | 0.82 |
| We should standardize which LLMs clinicians can use for decision-making support | 3.94 | 1.11 |
| If I use an LLM, I always make sure it cites a source and I assess the validity of the source independently | 3.77 | 1.44 |
| I spend significant time checking the accuracy of LLM-generated answers for clinical decision-making | 3.69 | 1.02 |
| My current institution has a clear formal or informal stance on the use of LLMs for clinical purposes | 2.95 | 1.22 |
| Concerns about energy and water consumption influence the likelihood or frequency of my AI/LLM use | 2.95 | 1.27 |
| LLMs should not be used to write notes or other clinical documentation | 2.31 | 0.98 |

\* 1 = 'strongly disagree' ; 5 = 'strongly agree'

#### Qualitative Findings from Journal Article Discussion Sessions

In total, 21 structured note-taking documents were collected from approximately 60 in-person conference attendees, and another two were submitted via the virtual form (N=23). Participants emphasized that use of LLMs must consider six areas of importance: Value and Efficiency Gains from AI, Deskilling and Never-Skilling, Governance and Oversight, Accuracy and AI Hallucination, Patient Safety and Education on AI Use.

##### Value and Efficiency Gains with AI

Participants expressed enthusiasm for potential value and efficiency gains from LLMs. This included widening the scope of previously infeasible research questions, particularly lengthy systematic reviews that require the synthesis of large volumes of detailed information. Others cited efficiency gains in clinical practice, such as in the “summary of clinical encounters” and how ambient listening-driven note generation can be “helpful and increase efficiency” (participant 2) in capturing critical information for writing time-intensive discharge summaries. Other participants noted, however, that LLMs are only helpful in these domains if the model is provided proper inputs; furthermore, they noted that humans are always responsible for how they use the final output.

##### Deskilling and Never-Skilling Due to AI

Participants’ responses generally focused on the idea that LLMs can be useful for augmentation of education and clinical practice but must not be used as a replacement for autonomous decision-making. They expressed concern that these technologies will reduce baseline learner curiosity, not necessarily due to laziness but rather due to its convenience amidst the rigor of medical training. For example, one participant suggested that the use of LLMs might lead to the “loss of one’s own ability to make decisions…and the skill of making decisions” (participant 7). Other participants wondered if medical education and some of the collaborative nature of clinical practice may suffer from a lack of differing perspectives if LLMs only express certain outputs.

##### Insufficient Governance and Oversight for AI

Lack of any sort of governance or oversight body was a salient concern among participants during discussion of LLM integration into the formation of clinical guidance. Participants acknowledged that the speed of AI development would make it extremely difficult to develop an effective oversight body, with some even noting that it is already too late to responsibly instill guardrails against its use. Other participants advocated governance frameworks, principles for responsibility and structure of LLM-driven guidelines, and clear institutional guidelines or policies for AI use. One participant asserted that governance structures are integral for “keeping a human in the loop for continued evaluation and monitoring” of LLM outputs, and that medicine may be able to “learn from other industries who are further along” in this effort (participant 10). Others focused on liability implications and identifying who might be liable for downstream results of misinformed LLM outputs, with one participant even suggesting lobbying the American Medical Association for regulation.

##### Assessing Accuracy and Identifying AI Hallucinations

Participants described concerns that LLMs do not admit they are wrong when they are indeed wrong, which could lead them to “doubling down” on detailed but incorrect treatment plans. They noted the need to “trust but verify” any output from LLMs and highlighted that this is particularly important in rapidly evolving, high-stakes medical specialties such as trauma, prehospital emergency care, and acute care. One participant also highlighted that clinicians must have the “quick ability to pivot” away from inaccurate outputs (participant 16). Some other participants were optimistic that LLM accuracy should continue to improve over time, but most noted that poor, ambiguous inputs into LLMs would lead to equally poor, ambiguous outputs that may cloud clinical judgment or entirely misinform novel learners. One participant suggested that when assessing AI accuracy in scenarios with significant ambiguity, the outputs should be compared to what a typical current physician would do rather than the “correct” or gold standard answer.

##### AI Implications for Patient Safety

Participants acknowledged that LLMs are “here to stay” and noted that their clinical implementation would impact patient safety. No one suggested that LLMs could augment patient safety. Some participants expressed a desire to better understand the audience for LLM tools and what they are intended to do, while others stated that AI is fundamentally built for a different purpose than clinical practice. Few participants mentioned the ethical considerations of LLM use, since outputs have difficulty incorporating patient priorities in their care and do not consult humans who may have an evolving understanding of a clinical scenario. Furthermore, one respondent expressed concern that hospital use of AI may prioritize monetary value over patient safety. Other participants focused on patient use of AI, noting that these tools have made many patients confused about what information (and which sources of information) to trust. To this point, one participant noted that LLMs are susceptible to “trash in/trash out in the information they give,” and that since patients may not know what information to give the LLM, it can lead to incorrect, incomplete, or even dangerous medical advice (participant 23).

##### Need for Clinical Education on AI Use

Participants also focused on LLMs’ inability to say “I don’t know” as a human might when generating an output. They noted that for this reason, clinicians and learners need further education on how to discern quality AI-generated answers from hallucinations or contextually inaccurate information. One stated that this is critical because “the principal responsibility of all decision making must remain with the physician” (participant 3).

Others focused on the bias that LLMs may contain based on the inputs they have been trained on; some cited concerns they hear frequently about how this may lead to biased outputs.^27^ Several participants supported providing formal training for medical students and residents to promote LLMs as an effective adjunct in practice, with one even calling for this training to be explicitly built into medical school curricula. Another participant suggested that other industries may have useful playbooks for AI adoption.

#### Post-Conference Survey Data

The majority (96%) of post-conference survey respondents (n=52) either somewhat or strongly agreed that clinical training curricula should include learning about effective LLM use, and that further education is essential for safely integrating LLMs into clinical decision-making. In addition, lack of knowledge on how to appropriately prompt LLMs was identified as one of three main inhibitors respondents cited regarding their own LLM use in clinical decision-making. Time required to verify accuracy of LLM outputs and concerns about patient harm were the other two frequently identified inhibitors. Table 3 shows all post-conference barriers that attendees cited to LLM use (on a scale of 1 = ‘not at all’ up to 3 = ‘very’).

**Table 3.** Inhibitors to LLM Use in Clinical Decision-Making (N=52 unless noted.

|  | **Mean** | **SD** |
| --- | --- | --- |
| Time required to verify accuracy | 2.37 | 0.69 |
| Concern about patient harm | 2.33 | 0.62 |
| Lack of personal training/knowledge on appropriate prompting | 2.21 | 0.80 |
| Legal/liability concerns | 2.12 | 0.65 |
| Institutional policies (N=49) | 2.02 | 0.69 |
| Energy/water consumption concerns | 1.79 | 0.72 |

**\*Given de-identified nature of post-conference survey, responses not limited to clinicians**

Finally, nearly two thirds of post-conference survey respondents indicated they would be likely to use LLMs to review existing screening or treatment guidelines. Over half indicated they would likely use them to review medication adverse effects, differential diagnoses, and choosing the proper medication. They reported being least likely to use LLMs for interpretating lab values or calculating medication dosing. Table 4 shows likelihood of LLM use by clinical scenario.

**Table 4.** Likelihood of LLM use by Clinical Scenario (N=52)*

| **What is the likelihood of using LLMs in the following instances?** |  |  |  |  |
| --- | --- | --- | --- | --- |
| **Survey Item** | **Likely** |  | **Unlikely** |  |
|  | **n** | **%** | **n** | **%** |
| Review of existing guidelines for screening or treatment protocols | 33 | 63.5 | 19 | 36.5 |
| Review of medication adverse side effects | 30 | 57.7 | 22 | 42.3 |
| Differential diagnoses / clinical workup | 27 | 51.9 | 25 | 48.1 |
| Medication choice | 27 | 51.9 | 25 | 48.1 |
| Research on off-label medication use or alternative treatment options | 25 | 48.1 | 27 | 51.9 |
| Dose calculations | 22 | 42.3 | 30 | 57.7 |
| Interpretation of lab values | 18 | 34.6 | 34 | 65.4 |

**\*Given de-identified nature of post-conference survey, responses not limited to clinicians.**

## Discussion

The rise of LLMs in medical education and clinical decision support, as well as the questions posed by its speed of adoption, was a highly engaging topic of conversation at the third DFI Conference. Group discussions and their data indicated excitement regarding ways that LLMs may enable new research capabilities, improve clinical efficiency, and become a useful adjunct resource for medical information. Many concerns, however, arose about these programs; namely, participants cited apprehension about the long-term impacts of AI and LLM use on the formation of clinical skills in medical training and maintenance of these skills in clinicians who trained before these programs existed. They also noted possible patient safety issues given these programs’ frequent inaccuracies. Attendees largely expressed that certain actions may help harness the useful components of AI while minimizing some possible negative effects. These included adding additional layers of governance and accountability to AI use, which may be most impactful if they are from the institutional or even governmental level. They also called for the integration of education on LLM use into medical school curricula as well as for currently practicing physicians and other medical practitioners. These recommendations, among others, are further detailed in the Rehani, et al (2026) paper of this supplement.

### Limitations

Structured note-taking documents were the main source of data used to identify participants’ key conference takeaways regarding LLM use, but they were only collected from participants who voluntarily agreed to both use the template and submit their notes. Those who submitted their notes may have felt most strongly about these issues. Each data set was independent, so it cannot be assumed that the quantitative and qualitative findings represent the same individuals, nor can they be directly linked. Furthermore, as in the two previous DFI conferences, attendees were predominantly from urban US academic programs who are engaged in national clinical guidance development. Prehospital clinicians (e.g., paramedics, emergency medical technicians, and EMS physicians) were not captured as a discrete provider type, limiting generalizability to the prehospital phase of trauma care, where clinical guidance such as field triage criteria is operationalized. Selection bias of attendees at the conference may be a limitation. Given that this was the second consecutive DFI conference featuring AI as a focus area, it is possible that participants were primed or influenced by DFI #2 content in a way that impacted their responses or willingness to respond in year three.^28,29^

### Next Steps

Attendees agreed that AI and LLM adoption in medical education and clinical practice have progressed more rapidly than any regulatory body or institution has been able to respond. Reasonable suggestions to provide training on higher-quality LLMs emerged from conference discussions and data. To provide optimal training, further research should first identify frequent use cases of LLMs across trainees and practicing clinicians. This would help tailor educational efforts more effectively and possibly enable health organizations to develop policies that more clearly delineate criteria for acceptable and unacceptable LLM use. Future work should also examine LLM use in time-critical and resource-constrained settings, including prehospital care and mass casualty or disaster response, where the time required to verify outputs (the most highly rated barrier in this study) is least available and where network or system downtime may limit access altogether.

Further, AI is largely trained on models, care, and clinical trials in the United States, particularly urban academic centers. This was a frequently cited concern in informal conversations at the conference and is frequently cited in the literature.^30^ Future research should address this concern and identify ways in which LLMs can be more equitably representative of resource-diverse settings.

## Conclusion

Given the increasing influence of LLMs on clinical practice and health professionals’ education, their responsible integration requires attention to both potential benefits and associated risks. Participants in this study recognized opportunities for improved efficiency and information synthesis while expressing concerns regarding accuracy, patient safety, clinical skill development, and governance. Findings highlight the importance of preparing clinicians and learners to critically evaluate LLM-generated information and of developing clear organizational approaches to appropriate use, verification, and oversight. As LLM technologies continue to evolve, ongoing evaluation will be necessary to determine how these tools can augment clinical practice and education without compromising clinical reasoning, professional accountability, or patient safety.

## Data Availability

Deidentified quantitative data produced in the present study are available upon reasonable request to the corresponding author.

## ACKNOWLEDGEMENTS

The Design for Implementation: The Future of Trauma Research C Clinical Guidance (DFI) conference series was made possible, in part, by a conference grant from the Agency for Healthcare Research and Quality (1R13HS028940-01A1). The views expressed in written conference materials or publications and by speakers and moderators do not necessarily reflect the official policies of the Department of Health and Human Services; nor does mention of trade names, commercial practices, or organizations imply endorsement by the U.S. Government.

The American Association for the Surgery of Trauma, American Burn Association, American Trauma Society, American College of Emergency Physicians, American Society of Anesthesiologists, Eastern Association for the Surgery of Trauma, Chest Wall Injury Society, Orthopaedic Trauma Association, Pediatric Trauma Society, Society of Critical Care Medicine, Society of Trauma Nurses, and Trauma Center Association of America provided travel funds for representatives to attend the 2025 DFI meeting. The Society of Trauma Nurses provided additional financial support. The American College of Surgeons hosted the event in its Chicago offices and provided meeting management, facilities, and audiovisual equipment at no cost to the conference.

Publication of this supplement is made possible by Medical Center of the Rockies, UCHealth (Loveland, Colorado, United States). Medical Center of the Rockies did not have any role in determining the results of this manuscript.

---

Checklist for Reporting of Survey Studies (CROSS)
| Section/topic | Item | Item description | Reported on page # |
| --- | --- | --- | --- |
| **Title and abstract** |  |  |  |
| Title and abstract | 1a | State the word “survey” along with a commonly used term in title or abstract to introduce the study’s design. | 13 |
|  | 1b | Provide an informative summary in the abstract, covering background, objectives, methods, findings/results, interpretation/discussion, and conclusions. | 13 |
| **Introduction** |  |  |  |
| Background | 2 | Provide a background about the rationale of study, what has been previously done, and why this survey is needed. | 15-16 |
| Purpose/aim | 3 | Identify specific purposes, aims, goals, or objectives of the study. | 16 |
| **Methods** |  |  |  |
| Study design | 4 | Specify the study design in the methods section with a commonly used term (e.g., cross-sectional or longitudinal). | 17 |
|  | 5a | Describe the questionnaire (e.g., number of sections, number of questions, number and names of instruments used). | Supplemental Items 2 & 3 |
| Data collection methods | 5b | Describe all questionnaire instruments that were used in the survey to measure particular concepts. Report target population, reported validity and reliability information, scoring/classification procedure, and reference links (if any). | 17 |
|  | 5c | Provide information on pretesting of the questionnaire, if performed (in the article or in an online supplement). Report the method of pretesting, number of times questionnaire was pre-tested, number and demographics of participants used for pretesting, and the level of similarity of demographics between pre-testing participants and sample population. | 18 |
|  | 5d | Questionnaire if possible, should be fully provided (in the article, or as appendices or as an online supplement). | Supple-mental Items 2 & 3 |
| Sample characteristics | 6a | Describe the study population (i.e., background, locations, eligibility criteria for participant inclusion in survey, exclusion criteria). | 17 |
|  | 6b | Describe the sampling techniques used (e.g., single stage or multistage sampling, simple random sampling, stratified sampling, cluster sampling, convenience sampling). Specify the locations of sample participants whenever clustered sampling was applied. | 17-18 |
|  | 6c | Provide information on sample size, along with details of sample size calculation. | 20 |
|  | 6d | Describe how representative the sample is of the study population (or target population if possible), particularly for population-based surveys. | n/a |
| Survey administration | 7a | Provide information on modes of questionnaire administration, including the type and number of contacts, the location where the survey was conducted (e.g., outpatient room or by use of online tools, such as SurveyMonkey). | 17-18 |
|  | 7b | Provide information of survey's time frame, such as periods of recruitment, exposure, and follow-up days. | 17 |
|  | 7c | Provide information on the entry process:<br>→For non-web-based surveys, provide approaches to minimize human error in data entry.<br>→For web-based surveys, provide approaches to prevent "multiple participation" of participants. | 17-19 |
| Study preparation | 8 | Describe any preparation process before conducting the survey (e.g., interviewers' training process, advertising the survey). | N/A |
| Ethical considerations | 9a | Provide information on ethical approval for the survey if obtained, including informed consent, institutional review board [IRB] approval, Helsinki declaration, and good clinical practice [GCP] declaration (as appropriate). | 16 |
|  | 9b | Provide information about survey anonymity and confidentiality and describe what mechanisms were used to protect unauthorized access. | 17 |
| Statistical analysis | 10a | Describe statistical methods and analytical approach. Report the statistical software that was used for data analysis. | 18-19 |
|  | 10b | Report any modification of variables used in the analysis, along with reference (if available). | N/A |
|  | 10c | Report details about how missing data was handled. Include rate of missing items, missing data mechanism (i.e., missing completely at random [MCAR], missing at random [MAR] or missing not at random [MNAR]) and methods used to deal with missing data (e.g., multiple imputation). | N/A |
|  | 10d | State how non-response error was addressed. | N/A |
|  | 10e | For longitudinal surveys, state how loss to follow-up was addressed. | N/A |
|  | 10f | Indicate whether any methods such as weighting of items or propensity scores have been used to adjust for non-representativeness of the sample. | N/A |
|  | 10g | Describe any sensitivity analysis conducted. | N/A |
| **Results** |  |  |  |
| Respondent characteristics | 11a | Report numbers of individuals at each stage of the study. Consider using a flow diagram, if possible. | 20 |
|  | 11b | Provide reasons for non-participation at each stage, if possible. | N/A |
|  | 11c | Report response rate, present the definition of response rate or the formula used to calculate response rate. | 20; cannot determine exact rate |
|  | 11d | Provide information to define how unique visitors are determined. Report number of unique visitors along with relevant proportions (e.g., view proportion, participation proportion, completion proportion). | N/A |
| Descriptive results | 12 | Provide characteristics of study participants, as well as information on potential confounders and assessed outcomes. | 20 |
| Main findings | 13a | Give unadjusted estimates and, if applicable, confounder-adjusted estimates along with 95% confidence intervals and p-values. | N/A |
|  | 13b | For multivariable analysis, provide information on the model building process, model fit statistics, and model assumptions (as appropriate). | N/A |
|  | 13c | Provide details about any sensitivity analysis performed. If there are considerable amount of missing data, report sensitivity analyses comparing the results of complete cases with that of the imputed dataset (if possible). | N/A |
| **Discussion** |  |  |  |
| Limitations | 14 | Discuss the limitations of the study, considering sources of potential biases and imprecisions, such as non-representativeness of sample, study design, important uncontrolled confounders. | 26 |
| Interpretations | 15 | Give a cautious overall interpretation of results, based on potential biases and imprecisions and suggest areas for future research. | 27 |
| Generalizability | 16 | Discuss the external validity of the results. | 26-27 |
| **Other sections** |  |  |  |
| Role of funding source | 17 | State whether any funding organization has had any roles in the survey's design, implementation, and analysis. | 11 |
| Conflict of interest | 18 | Declare any potential conflict of interest. | 11 |
| Acknowledgements | 19 | Provide names of organizations/persons that are acknowledged along with their contribution to the research. | 11 |
| Sharma A, Minh Duc NT, Luu Lam Thang T, Nam NH, Ng SJ, Abbas KS, Huy NT, Marušić A, Paul CL, Kwok J, et al. A Consensus-Based Checklist for Reporting of Survey Studies (CROSS). *J Gen Intern Med*. 2021;36(10):3179-3187. DOI: 10.1007/s11606-021-06737-1. |  |  |  |

## Supplemental Items

Supplemental Item 1. Consensus-Based Checklist for Reporting of Survey Studies

Supplemental Item 2. Relevant Pre-Conference Survey Questions

Supplemental Item 3. Relevant Post-Conference Survey Questions

Supplemental Item 2. Relevant Pre-Conference Survey Questions

**3. In what country do you primarily live and work?**

- The United States of America
- Other

**4. To which gender identity do you most identify? [select one]**

- Woman
- Man
- Transgender
- Non-binary/non-conforming
- Other
- Prefer not to say

**5. What age category do you fall in? [select one]**

- Under 25
- 25-44
- 45-64
- 65 and older
- Prefer not to answer

**6. Are you Hispanic, Latino or of Spanish origin?**

- Yes
- No
- Prefer no to answer

**7. How would you describe your ethnicity? [select all that apply]**

- American Indian or Alaska Native
- Asian
- Black or African American
- Native Hawaiian or Other Pacific Islander
- White
- Prefer not to answer
- Other

**8. What educational degree(s) do you hold? [Select all that apply]**

- None
- Bachelor of Science and/or Arts (BS/BA)
- Bachelor of Science in Nursing (BSN)
- Master of Science and/or Arts (MS/MA)
- Master Of Science in Nursing (MSN)
- Master of Business Administration (MBA)
- Master of Public Health (MPH)
- Doctor of Medicine (MD)
- Doctor of Philosophy (PhD)
- Other

**9. Which medical professional societies/organizations are you affiliated with? (Select all that apply)**

- None / Not applicable
- American Association for the Surgery of Trauma (AAST)
- American Association of Neurological Surgeons (AANS)
- American Burn Association (ABA)
- American College of Emergency Physicians (ACEP)
- American College of Surgeons Committee on Trauma (ACS COT)
- American College of Surgeons (ACS)
- Military Health System Strategic Partnership
- American Geriatrics Society (AGS)
- American Trauma Society (ATS)
- Anticoagulation Forum
- Association of Out Surgeons and Allies (AOSA)
- Association of Women Surgeons (AWS)
- Congress of Neurological Surgeons
- Eastern Association for the Surgery of Trauma (EAST)
- Latino Surgical Society (LSS)
- National Association Of Emergency Medicine Technicians (NAEMT)
- National Blood Clot Alliance
- North American Thrombosis Forum
- Orthopaedic Trauma Association (OTA)
- Society of Asian American Surgeons (SAAS)
- Society Of Critical Care Medicine (SCCM)
- Society of Trauma Nurses (STN)
- The American Society of Emergency Radiology
- The Health Alliance for Violence Intervention (HAVI)
- Trauma Center Association of America (TCAA)
- Western Trauma Association (WTA)
- Other

**10. Do you consider where you primarily live and work to be rural?**

- Yes
- No
- Not applicable

**11. Do you have a current and/or prior role in the military?**

- Yes
- No
- Not applicable

**12. Do you consider yourself to be a patient or patient advocate?**

- Yes
- No
- Not applicable

**13. Do you hold a professional role in diversity, equity, and inclusion (DEI) initiatives?**

- Yes
- No
- Not applicable

**14. Are you directly involved in global health initiatives?**

- Yes
- No
- Not applicable

**15. What provider type are you?**

- N/A / Not a clinician
- Physician
- APP
- RN
- EMT
- Other

**16. Do you treat pediatric patients in the course of your clinical duties?**

- Yes
- No
- Not applicable

**17. What is your clinical discipline? (Select all that apply)**

- None / Not Applicable
- Trauma Surgery
- General Surgery
- Pediatrics
- Pulmonary
- Burns
- Orthopaedics
- Emergency Medicine
- Vascular
- Neurology
- Anesthesia
- Physical and Rehabilitation Medicine
- Neurosurgery
- Internal Medicine
- Family Medicine
- Radiology
- Other

**18. Do you currently work in a verified/designated trauma center?**

- Yes
- No
- Not applicable

**19. If yes, what is the trauma center’s level?**

- Level I
- Level II
- Level III
- Level IV
- Level V
- Other

**20. Is participating in dissemination science, or quality improvement, a core part of your job description?**

- Yes
- No
- Not applicable

**21. Is conducting research a core part of your job description?**

- Yes
- No
- Not applicable

**22. Do you participate in clinical guidance development?**

- Yes
- No
- Not applicable

**23. If yes, in what context? (Select all that apply)**

- International
- National
- Regional
- Institutional
- Other

**24. How are you planning on attending the conference?**

- In-person
- Remotely (via Zoom)

## Participant Guidance

*If you are a non-clinician, please answer according to your own health-information seeking practices. For questions with stems that include ’should’, answer according to what you wish/think clinicians should be doing*.

**25. When you face clinical uncertainty, what resource do you turn to most often? [select one]**

- Peers and peer networks
- Peer-reviewed published literature
- Summative texts and point-of care resources (e.g. UpToDate)
- Electronic health recorded integrated tool
- LLMs or AI
- Other

**26. How often do you use artificial intelligence (AI) or large language models (LLMs, i.e. ChatGPT, OpenEvidence, Claude, etc.) for support in clinical decision-making?**

- Daily
- Weekly
- less than monthly
- Monthly
- Never

**27. In what proportion of clinical encounters do you use AI / an LLM? (e.g. if you see 20 patients on rounds, in how many of those patient encounters would you use an LLM?)**

- NA/None
- Most
- About half
- A minority
- Very few

**28. If you use LLMs, what platform are you most likely to use?**

- ChatGPT
- OpenEvidence
- Gemini
- Claude
- Llama
- Other

**29. Please tell us more about why you have been using your preferred platform(s).**

*Enter your answer*

**30. How often do you see others using artificial intelligence (AI) or learned language models (LLMs) (e.g. ChatGPT, OpenEvidence, Claude) for support in clinical decision-making?**

- Not at all
- Rarely
- Sometimes
- Often
- Regularly
- **31. What platform do you see them use most commonly?**
- ChatGPT
- OpenEvidence
- Gemini
- Claude
- Llama
- Other

**32. Whether you use LLMs yourself or not, are there any situations or use cases where you consider LLMs to be particularly useful? (select all that apply)**

- I do not find LLMs to be useful
- Differential diagnoses / clinical workup
- Medication choice
- Dose calculation
- Interpretation of lab values
- Review of medication side effects (ASE)
- Review of existing guidelines for screening or treatment protocols
- Research on off-label medication use or alternative treatment options
- Other

**33. Whether you use LLMs yourself or not, are there any situations or use cases where you consider LLMs to be particularly dangerous or unhelpful? (select all that apply)**

- I do not find LLMs to be dangerous or unhelpful
- Differential diagnoses / clinical workup
- Medication choice
- Dose calculation
- Interpretation of lab values
- Review of medication side effects (ASE)
- Review of existing guidelines for screening or treatment protocols
- Research on off-label medication use or alternative treatment options
- Other

**34. If I use an LLM in any clinical decision-making capacity, I always make sure it cites a source, and I assess the validity of the source independently.**

- Strongly disagree
- Somewhat disagree
- Neutral
- Somewhat agree
- Strongly agree

**35. I have to spend significant time checking the accuracy of answers generated by LLMs as it pertains to clinical decision-making.**

- Strongly disagree
- Somewhat disagree
- Neutral
- Somewhat agree
- Strongly agree

**36. My current institution has a clear formal or informal stance on the use of LLMs for clinical purposes.**

- NA / I’m not clinically employed
- Strongly disagree
- Somewhat disagree
- Neutral
- Somewhat agree
- Strongly agree

**37. Clinical training curriculums should include learning how to use LLMs effectively.**

- Strongly disagree
- Somewhat disagree
- Neutral
- Somewhat agree
- Strongly agree

**38. We should standardize which LLMs clinicians can use for decision-making support.**

- Strongly disagree
- Somewhat disagree
- Neutral
- Somewhat agree
- Strongly agree

**39. Concerns about energy and water consumption have an influence on the likelihood of using AI/LLMs in clinical decision-making or the frequency with which I would use them.**

- Strongly disagree
- Somewhat disagree
- Neutral
- Somewhat agree
- Strongly agree

**40. LLMs should not be used to write notes or other clinical documentation.**

- Strongly disagree
- Somewhat disagree
- Neutral
- Somewhat agree
- Strongly agree

**41. LLMs used to support clinical decisions should be regularly audited for quality control.**

- Strongly disagree
- Somewhat disagree
- Neutral
- Somewhat agree
- Strongly agree

**42. Have you ever experienced a clinical situation in which you found that LLM use significantly improved your / a colleague’s / a trainee’s clinical decision-making?**

- No
- Yes

**43. If yes, please elaborate:**

- *Enter your answer*

**44. Have you ever experienced a clinical situation in which you found that LLM use was significantly detrimental to your / a colleague’s / a trainee’s clinical decision-making?**

- No
- Yes
- **45. If yes, please elaborate:**

*Enter your answer*

## Supplemental Item 3. Relevant Post-Conference Survey Questions

**17. Does the frequency of LLM use have an adverse climate impact?**

- No
- Unsure or do not know
- Somewhat agree
- Yes, but minimal
- Yes, a significant impact
- Other

**18. I have concerns about the rise of LLM use in clinical decision-making and the potential harmful impacts it may have.**

- Strongly disagree
- Somewhat disagree
- Neutral
- Somewhat agree
- Strongly agree

**19. How significant are each of the below as a barrier to your current or potential use of LLMs in clinical decision-making?**

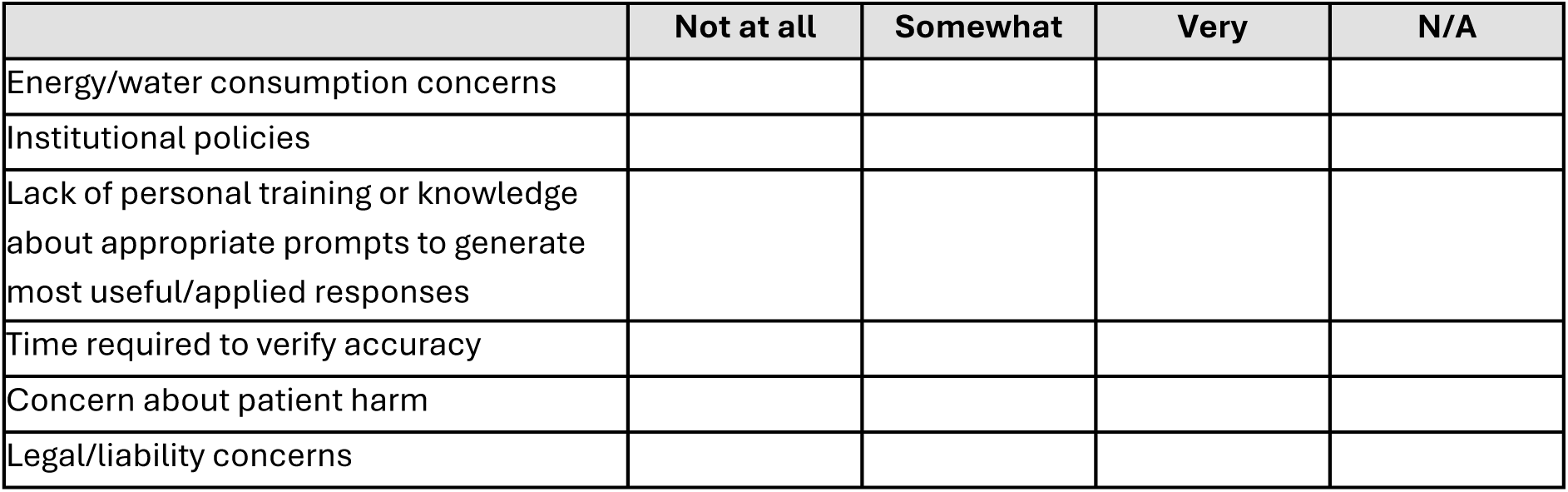

- **20. If you can think of other potential barriers, please elaborate:**

*Enter your answer*

**21. What is the likelihood of your using LLMs in the following instances:**

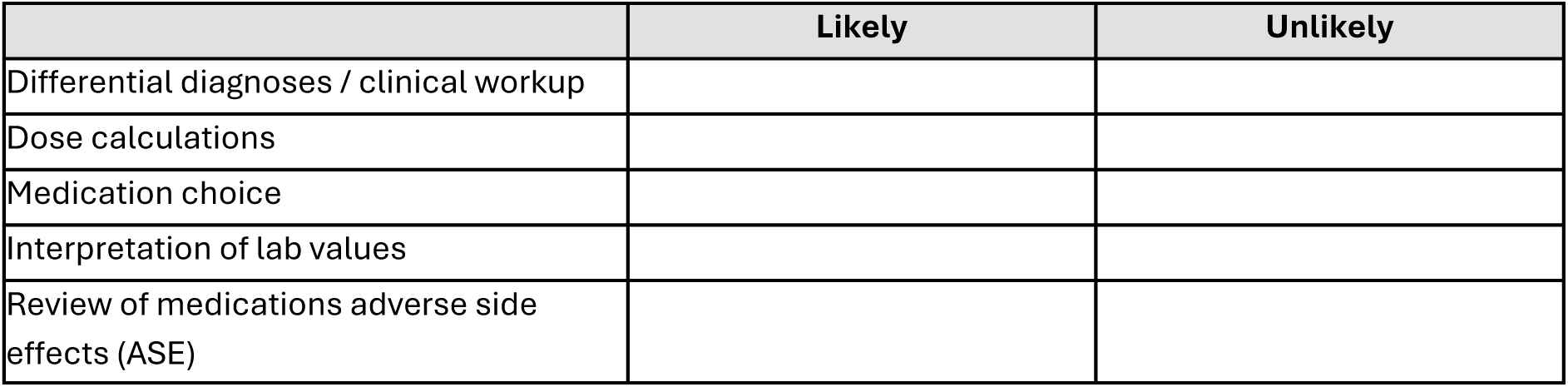
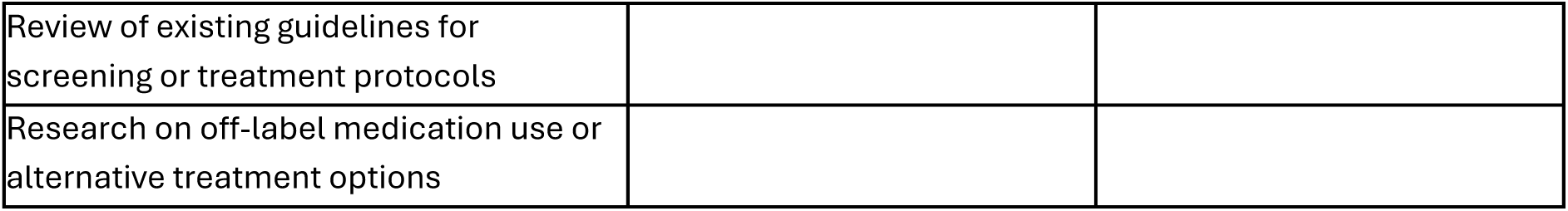

- **22. If there are other instances or scenarios you think are important to address, please elaborate below:**

*Enter your answer*

**23. Clinical training curricula should include learning how to use LLMs effectively.**

- Strongly disagree
- Somewhat disagree
- Neutral
- Somewhat agree
- Strongly agree

**24. For LLMs to be more safely integrated into clinical decision-making, further education is needed regarding how and when to use them.**

- Strongly disagree
- Somewhat disagree
- Neutral
- Somewhat agree
- Strongly agree

**25. Please elaborate on your answer above, and provide any additional context you feel is important.**

*Enter your answer*

## ETHICS APPROVAL

This study involves human participants. This study was reviewed by the Colorado Multiple Institutional Review Board, CB F490; COMIRB No: 24-0047, and determined exempt from institutional review board review because it was deemed category 2. Category 2 reads, “Research that only includes interactions involving educational tests (cognitive, diagnostic, aptitude, achievement), survey procedures, interview procedures, or observation of public behavior (including visual or auditory recording) if at least one of the following criteria is met: (A) the information obtained is recorded by the investigator in such a manner that the identity of the human subjects cannot readily be ascertained, directly or through identifiers linked to the subjects; (B) any disclosure of the human subjects’ responses outside the research would not reasonably place the subjects at risk of criminal or civil liability or be damaging to the subjects’ financial standing, employability, educational advancement, or reputation; or (C) the information obtained is recorded by the investigator in such a manner that the identity of the human subjects can readily be ascertained, directly or through identifiers linked to the subjects, and an IRB conducts a limited IRB review to make the determination required by 45 CFR 46.111(a)(7).” Participants gave informed consent to participate in the study before taking part.

## AUTHOR CONTRIBUTIONS

Design: BF, LNL, HG, AN, MA

Data acquisition: BF, CR, EC, SE, LNL, LV, AR, SR, CC, RR, KG, AM

Analysis and interpretation: BF, KS, AM

Drafting and Critical Revision: BF, AM, LNL, AN, 2026 Design for Implementation (DFI) Authorship Group

Guarantor: LNL

## References

1. Shy, B. D., Baloescu, C., Faustino, I. V., Taylor, R. A., Gottlieb, M., Sangal, R. B., Hood, C., Genes, N., Rabin, E. J., C ACEP AI Task Force (2025). Early Insights Among Emergency Medicine Physicians on Artificial Intelligence: A National, Convenience-sample Survey of the American College of Emergency Physicians. Journal of the American College of Emergency Physicians open, 7(1), 100308. 10.1016/j.acepjo.2025.100308

2. Soltani, T., Kim, P., McKinley, W. I., Frasier, L., Dowzicky, P., Holena, D. N., Callcut, R. A., C Eastern Association for the Surgery of Trauma’s Quality, Safety, and Outcomes Committee (2026). Evaluation framework for assessing the quality and safety of artificial intelligence solutions for the acute care surgeon: a narrative review from the Eastern Association for the Surgery of Trauma quality, safety, and outcomes committee. Trauma surgery C acute care open, 11(3), e002163. 10.1136/tsaco-2025-002163

3. Abbasi J, Hswen Y. Blind Spots, Shortcuts, and Automation Bias—Researchers Are Aiming to Improve AI Clinical Models. JAMA. 2024;331(11):903–906. doi:10.1001/jama.2023.28262

4. Yang, R., Ning, Y., Keppo, E., Liu, M., Hong, C., Bitterman, D. S., Ong, J. C. L., Ting, D. S. W., C Liu, N. (2025). Retrieval-augmented generation for generative artificial intelligence in health care. npj Health Systems, 2, Article 2. 10.1038/s44401-024-00004-1

5. Kung TH, Cheatham M, Medenilla A, et al. Performance of ChatGPT on USMLE: Potential for AI-assisted Medical Education Using Large Language Models. PLOS Digital Health. 2023;2(2):e0000198. doi:10.1371/journal.pdig.0000198

6. OpenEvidence. (2025, August 15). OpenEvidence creates the first AI in history to score a perfect 100% on the United States Medical Licensing Examination (USMLE). OpenEvidence. https://www.openevidence.com/announcements/openevidence-creates-the-first-ai-in-history-to-score-a-perfect-100percent-on-the-united-states-medical-licensing-examination-usmle

7. OpenEvidence. (n.d.). OpenEvidence: America’s official medical knowledge platform. https://www.openevidence.com

8. Ke, Y., Jin, L., Ong, J. C. L., Thirunavukarasu, A. J., Car, J., Cheung, C. Y., Tham, Y. C., Ting, D. S. W., Ong, M. E. H., Compton, S., Narayan, A., Keane, P. A., Wong, T. Y., Bates, D. W., Tan, P., C Liu, N. (2026). AI-induced never-skilling in medical education. Nature medicine, 10.1038/s41591-026-04438-y. Advance online publication. https://doi-org.cuanschutz.idm.oclc.org/10.1038/s41591-026-04438-y

9. Monteith, S., Glenn, T., Geddes, J. R., Whybrow, P. C., Achtyes, E. D., Bauer, R., C Bauer, M. (2026). Artificial intelligence and deskilling in medicine. The British journal of psychiatry : the journal of mental science, 1–3. Advance online publication. 10.1192/bjp.2025.10496

10. Keren, R., Desai, B. R., C West, D. C. (2026). Promoting Clinical Expertise in the Age of AI: No Struggle, No Mastery. JAMA, 335(21), 1837–1838. 10.1001/jama.2026.6097

11. Pazzaglia, E., Violante, T., Rottoli, M., C Raimondo, D. (2026). Laparoscopic Deskilling-The Hidden Risk of Robotic Dominance. JAMA surgery, 161(9), 859–860. 10.1001/jamasurg.2026.3048

12. Budzyń, K., Romańczyk, M., Kitala, D., Kołodziej, P., Bugajski, M., Adami, H. O., Blom, J., Buszkiewicz, M., Halvorsen, N., Hassan, C., Romańczyk, T., Holme, Ø., Jarus, K., Fielding, S., Kunar, M., Pellise, M., Pilonis, N., Kamiński, M. F., Kalager, M., Bretthauer, M., … Mori, Y. (2025). Endoscopist deskilling risk after exposure to artificial intelligence in colonoscopy: a multicentre, observational study. The lancet. Gastroenterology C hepatology, 10(10), 896–903. 10.1016/S2468-1253(25)00133-5

13. Vishwanath, K., Alyakin, A., Ghosh, M. et al. General-purpose large language models outperform specialized clinical AI tools on medical benchmarks. Nat Med 32, 2405–2409 (2026). 10.1038/s41591-026-04431-5

14. Wu, D., Haredasht, F. N., Maharaj, S. K., Jain, P., Tran, J., Gwiazdon, M., Rustagi, A., Jindal, J., Koshy, J. M., Kadiyala, V., Agarwal, A., Tappuni, B., French, B., Jesudasen, S., Cosgriff, C. V., Chakraborty, R., Caldwell, J., Ziolkowski, S., Iberri, D. J., Diep, R., … Goh, E. (2025). First, do NOHARM: towards clinically safe large language models. ArXiv, arXiv:2512.01241v2.

15. Yang, X., Xiao, Y., Liu, D., Deng, H., Huang, J., Zhou, Y., Liang, M., Dong, L., Yuan, Z., Yao, J., Guo, W., C Xu, C. (2025). Factors Influencing Adoption of Large Language Models in Health Care: Multicenter Cross-Sectional Mixed Methods Observational Study. Journal of medical Internet research, 27, e84918. https://doi-org.cuanschutz.idm.oclc.org/10.2196/84918

16. Solmonovich, R. L., Kouba, I., Lee, J. Y., Demertzis, K., C Blitz, M. J. (2025). Physician awareness of, interest in, and current use of artificial intelligence large language model-based virtual assistants. PloS one, 20(5), e0320749. 10.1371/journal.pone.0320749

17. Wilson DJ, Gellings JA, Zavala G, Steiner A, Kwan BM, Bixby PJ, et al. Conference Proceedings for the 2024 Design for Implementation: The Future of Trauma Research and Clinical Guidance Conference Series. Trauma Surgery C Acute Care Open. 2025;10:e001583. 10.1136/tsaco-2024-001583

18. Wilson DJ, Gellings JA, Frankel B, Zavala Wong G, Punch L, Lang KL, et al. Conference proceedings for the 2025 Design for Implementation: The Future of Trauma Research and Clinical Guidance Conference Series. Trauma Surgery C Acute Care Open. 2025;10:e002059. 10.1136/tsaco-2025-002059

19. Sharma A, Minh Duc NT, Luu Lam Thang T, et al. A Consensus-Based Checklist for Reporting of Survey Studies (CROSS). J Gen Intern Med 2021; 36:3179–87. doi:10.1007/s11606-021-06737-1

20. Berzin, T. M., C Topol, E. J. (2025). Preserving clinical skills in the age of AI assistance. Lancet (London, England), 406(10513), 1719. https://doi-org.cuanschutz.idm.oclc.org/10.1016/S0140-6736(25)02075-6

21. Goh, E., Gallo, R., Hom, J., Strong, E., Weng, Y., Kerman, H., Cool, J. A., Kanjee, Z., Parsons, A. S., Ahuja, N., Horvitz, E., Yang, D., Milstein, A., Olson, A. P. J., Rodman, A., C Chen, J. H. (2024). Large Language Model Influence on Diagnostic Reasoning: A Randomized Clinical Trial. JAMA network open, 7(10), e2440969. https://doi-org.cuanschutz.idm.oclc.org/10.1001/jamanetworkopen.2024.40969

22. Khraisha, Q., Put, S., Kappenberg, J., Warraitch, A., C Hadfield, K. (2024). Can large language models replace humans in systematic reviews? Evaluating GPT-4’s efficacy in screening and extracting data from peer-reviewed and grey literature in multiple languages. Research synthesis methods, 15(4), 616–626. https://doi-org.cuanschutz.idm.oclc.org/10.1002/jrsm.1715

23. Bean, A.M., Payne, R.E., Parsons, G. et al. Reliability of LLMs as medical assistants for the general public: a randomized preregistered study. Nat Med 32, 609–615 (2026). 10.1038/s41591-025-04074-y

24. Kowalski, C. P., Nevedal, A. L., Finley, E. P., Young, J. P., Lewinski, A. A., Midboe, A. M., C Hamilton, A. B. (2024). Planning for and Assessing Rigor in Rapid Qualitative Analysis (PARRQA): a consensus-based framework for designing, conducting, and reporting. Implementation science : IS, 19(1), 71. https://doi-org.cuanschutz.idm.oclc.org/10.1186/s13012-024-01397-1

25. Hsieh, H. F., C Shannon, S. E. (2005). Three approaches to qualitative content analysis. Qualitative health research, 15(9), 1277–1288. 10.1177/1049732305276687

26. Nevedal, A. L., Reardon, C. M., Opra Widerquist, M. A., Jackson, G. L., Cutrona, S. L., White, B. S., C Damschroder, L. J. (2021). Rapid versus traditional qualitative analysis using the Consolidated Framework for Implementation Research (CFIR). Implementation science : IS, 16(1), 67. 10.1186/s13012-021-01111-5

27. Seale S, Shah A, Kundu S, Liebert CA. Gender Bias in AI-Generated Letters of Recommendation for Surgical Trainees. JAMA Surg. Published online September 23, 2026. doi:10.1001/jamasurg.2026.4156

28. Zavala Wong, G., Rosenauer, S., Church, C., Sherifali, D., Racey, M., Grider, K., Moreno, A. N., LaGrone, L., C 2025 Design for Implementation (DFI) Authorship Group (2026). Better way: initial acceptability testing of using artificial intelligence tools to accelerate development of trauma clinical guidance. Trauma surgery C acute care open, 10(Suppl), e002060. 10.1136/tsaco-2025-002060

29. Frankel, B., Beasley, L., Rosenauer, S., Church, C., Grider, K., Buchner, M., Francoeur, N., Zavala Wong, G., Moreno, A. N., LaGrone, L., C 2025 Design for Implementation (DFI) Authorship Group (2026). Usability and feasibility of a mobile application for real-time trauma care guidance: considerations for user adoption. Trauma surgery C acute care open, 10(Suppl 5), e002051. 10.1136/tsaco-2025-002051

30. Alderman, J. E., Palmer, J., Laws, E., McCradden, M. D., Ordish, J., Ghassemi, M., Pfohl, S. R., Rostamzadeh, N., Cole-Lewis, H., Glocker, B., Calvert, M., Pollard, T. J., Gill, J., Gath, J., Adebajo, A., Beng, J., Leung, C. H., Kuku, S., Farmer, L. A., Matin, R. N., … Liu, X. (2025). Tackling algorithmic bias and promoting transparency in health datasets: the STANDING Together consensus recommendations. The Lancet. Digital health, 7(1), e64–e88. 10.1016/S2589-7500(24)00224-3

